# Predictors of Early Neurodevelopmental Outcomes for Children with Congenital Heart Disease: A Report from the Cardiac Neurodevelopmental Outcome Collaborative

**DOI:** 10.1101/2025.03.03.25323281

**Authors:** Mike Seed, Dawn Ilardi, Valerie Rofeberg, Cynthia Ortinau, Caren Goldberg, Garrett Reichle, Justin Elhoff, Amy Jo Lisanti, Jennifer Butcher, Caitlin Rollins, Lauren Bush, Andrew Van Bergen, Shabnam Peyvandi, Emily Bucholz, Stephanie Cox, Lyla Hampton, Jacqueline Sanz, Sonia Monteiro, Shruti Tewar, Kiona Allen, Caroline Lee, Kristi Glotzbach, Nneka Alexander, Laurel Bear, Corinne Anton, Renee Sananes, Linh Ly, Gina Boucher, Kelly Wolfe, Lindsay Edwards, Elizabeth Willen, Alexander Tan, Christina Ortega, Erica Sood, Anjali Sadhwani, Kari Crawford Plant, Lauren Quigley, Jessica Pliego, Elizabeth Valles, Abbey Hines, David Wypij, Thomas Miller

## Abstract

**Background:** Neurodevelopmental impairments are common in children with congenital heart disease. The Cardiac Neurodevelopmental Outcome Collaborative and Pediatric Cardiac Critical Care Consortium registry linkage allows for the analysis of associations between neurodevelopmental, medical, and sociodemographic variables in a large contemporary cohort.

**Methods:** Children with congenital heart disease who required surgery with cardiopulmonary bypass at <12 months of age and completed a neurodevelopmental assessment between 11-30 months of age from 2019-2022 were included. Multivariable regression modeling was performed to identify differences in Cognitive, Language, and Motor standard scores from the Bayley Scales of Infant and Toddler Development-III/4 based on congenital cardiac diagnosis, clinical risk factors, and social drivers of health.

**Results:** Primary analyses included 942 assessments from 868 children completed at 25 sites. Across cardiac diagnostic groups, those with genetic diagnoses (n=116 assessments) scored >1 standard deviation lower on all Bayley indices than those without (*P*<0.001 for each). For those without genetic diagnoses, there were differences between cardiac diagnostic groups (*P*<0.001) in both Cognitive and Motor indices; participants with transposition of the great arteries exhibited the highest scores compared with other cardiac diagnoses. Lower birth weight, male sex, older age at initial surgery, longer hospital length of stay, more cardiac catheterizations, and lower primary caregiver education were independently associated with worse performance in all indices.

**Conclusions:** Findings from this multicenter cohort demonstrate variation in neurodevelopmental outcomes according to cardiac diagnosis. Regardless of cardiac diagnosis, the presence of a genetic diagnosis is associated with lower neurodevelopmental scores. Heterogeneous outcomes reinforce the importance of surveillance for all infants undergoing heart surgery in the first year of life.

**Clinical Perspective:** *What is new?:*

- While genetic diagnoses confer the highest risk of developmental delays and disorders in patients with CHD, cardiac diagnosis also impacts early neurodevelopmental outcomes in non-syndromic patients.
- In non-syndromic patients, those with transposition of the great arteries exhibit higher scores on early ND testing than other common CHD subtypes, while those with single ventricle physiology and atrioventricular septal defects exhibit lower scores.
- Older age at surgery and greater number of interventional cardiac catheterizations may represent newly identified risk factors for adverse early ND outcomes in infants with CHD.

*What are the clinical implications?:*

- Data from the CNOC registry emphasizes the importance of ND follow-up for all infants undergoing cardiac surgery, including those with simpler CHD subtypes.
- The improved outcomes we observed in patients with transposition of the great arteries suggest advances in routine clinical management, including early surgery, may have had a neuroprotective influence.

## Introduction

Neurodevelopmental (ND) delays and disorders affect up to half of children requiring surgical intervention for congenital heart disease (CHD) during infancy.^1^ Early motor and language delays may progress to cognitive, academic, social-emotional, and executive functioning concerns at school age.^2^ Adults with CHD exhibit lower educational attainment, more unemployment, a higher incidence of mental health issues, and reduced quality of life.^3,4^ The etiology of ND differences in CHD is not fully understood and is likely multifactorial. Innate and prenatal factors such as genetic variation, maternal health, and fetal cardiovascular physiology combine with postnatal medical, surgical, and environmental risks to affect brain health and associated neurodevelopment.^5–7^ Specifically, autopsy and neuroimaging studies suggest the neurobiology generally reflects both abnormal brain development and acquired brain injury.

Observational studies have identified clinical risk factors predisposing to ND delay in CHD, including genetic diagnoses, prolonged hospitalization, and preterm birth.^2,10,11^ The severity of ND impairments has been linked to the complexity of the heart disease.^12,13^ Social drivers of health including maternal education and socioeconomic status are also associated with ND outcomes in children with CHD.^8,9^ Prior studies examining associations between risk factors and ND outcomes have typically grouped participants with a variety of cardiac diagnoses together or focused exclusively on a specific cardiac diagnosis. For example, a single center cohort of participants with transposition of the great arteries (D-TGA) has been studied extensively in the Boston Circulatory Arrest Trial,^14–17^ and ND outcomes for those with hypoplastic left heart syndrome (HLHS) have been reported from the multicenter Single Ventricle Reconstruction Trial.^18,19^ However, more recent studies suggest that associations between cardiac diagnosis and ND outcomes may be evolving as perioperative management changes, with D-TGA patients now exhibiting improved outcomes compared with earlier findings.^20^

The Cardiac Neurodevelopmental Outcome Collaborative (CNOC) collects ND outcome data in CHD patients through a multinational, multicenter clinical data registry.^21^ Partnership with the Pediatric Cardiac Critical Care Consortium (PC^4^), who collect a registry of detailed perioperative clinical information, has facilitated the integration of ND and perioperative data.^22^ CNOC site members participating in the registry use standardized ND measures. Assessment results are submitted through a web-based data platform (Arbormetrix), and data are managed and cleaned at the CNOC Data Coordinating Center located at the University of Michigan, audited by the Neurodevelopmental Core Lab at Children’s National Hospital and analyzed at the Data Analysis Core located at Boston Children’s Hospital. In tandem with the creation of the CNOC registry, a research committee prioritized two initial study aims. Findings from the first aim, to identify factors associated with attendance for cardiac ND assessment, were previously published.^22^ In the current study, we aimed to describe ND outcomes for a comprehensive sample of infants and toddlers undergoing repair during infancy to understand the medical risk factors and social drivers associated with ND scores. We hypothesized that ND outcomes would differ by cardiac diagnosis and that clinical risk factors and social drivers of health would be predictive of infant/toddler ND outcomes.

## Methods

### Study Design and Participants

The study was approved by the University of Michigan Institutional Review Board with appropriate data sharing agreements with each of the 27 CNOC sites that contributed data. The inclusion criteria required children to have undergone ND assessment with the Bayley Scales of Infant and Toddler Development-III/4 (Bayley) between 11 and 30 months of age from May 2019 to June 2022 following cardiopulmonary bypass surgery during the first year of life, the details of which needed to have been recorded in the PC^4^ registry.

### Sociodemographic and Medical Data

The CNOC and PC^4^ clinical data registries were used to collect sociodemographic and medical data. Sociodemographic variables included sex, race and ethnicity, primary caregiver education, and the Child Opportunity Index (COI), a metric of neighborhood resources and conditions based on the child’s zip code.^23^ Medical variables identified as pertinent by the cardiac ND literature included the presence of a genetic diagnosis, cardiac diagnosis, gestational age, birth weight, birth head circumference, age at first cardiopulmonary bypass surgery (index operation), type of surgical procedure(s), post-operative length of stay at the index operation, history of cardiac arrest, history of stroke or clinical or electrographic seizure, extracorporeal membrane oxygenation (ECMO) during the index operation, and number of cardiac operations and cardiac catheterizations in the first year of life. Cardiac diagnoses were divided amongst seven groups: dextro-transposition of the great arteries (D-TGA), coarctation of the aorta (CoA), ventricular septal defect (VSD), tetralogy of Fallot (TOF), single ventricle physiology (SVP), atrioventricular septal defect (AVSD), and other. Cardiac surgical procedures were classified according to the Society of Thoracic Surgeons-European Association for Cardio-Thoracic Surgery (STAT) system, with higher scores reflecting operations with higher risk of mortality.^24^

### Neurodevelopmental Data

One component of the ND assessment included the Bayley scales, which provide scores based on population-normed data.^25^ ND outcome variables included the Cognitive, Motor, and Language standard scores (population mean 100, standard deviation 15). During the study, we were notified that a conversion factor would need to be applied to scores obtained using version 4 of the Bayley. As the raw scores were required for conversion, we opted to exclude patients for whom Bayley 4 raw scores were not available and for whom there were scoring errors in our primary analyses. In addition, patients missing raw scores or with inconsistencies between raw and standard scores were only included in a separate sensitivity analysis.

### Statistical Analyses

Because of the known association of genetic disorders with adverse ND outcomes independent from CHD, patients were divided into those with versus without a genetic diagnosis. Graphical and regression methods were used to assess univariable associations and the impact of genetic diagnosis on ND outcomes. For those patients without a genetic diagnosis, stepwise multivariable regression modeling was performed to investigate associations between the ND outcomes for each index with medical variables based on a *P*-value <0.05. After controlling for cardiac diagnosis and Bayley version (III vs. 4), candidate predictors included sex and the medical characteristics listed above. To account for missing data for primary caregiver education and COI, these factors are added in individually with separate models. Throughout, robust variances from generalized estimating equation models with independence working correlations were used to control for center-to-center variability and repeat assessments from some patients. R version 4.3.0 (R Foundation for Statistical Computing, Vienna, Austria) was used for all analyses.

## Results

### Study Population and Characteristics

Figure 1 shows a flow diagram illustrating the final data available for analysis. Bayley scores and PC^4^ data were available for 1339 patients from 27 sites. After data validation, we had a total of 942 assessments representing 868 unique patients from 25 sites for primary analysis. Of these assessments, 116 had a genetic diagnosis while 826 had no genetic diagnosis. Thus, the final primary analysis dataset of patients without a genetic diagnosis available for analysis comprised 814 assessments for the Cognitive index, 764 for Language, and 791 for Motor.

**Figure 1.**
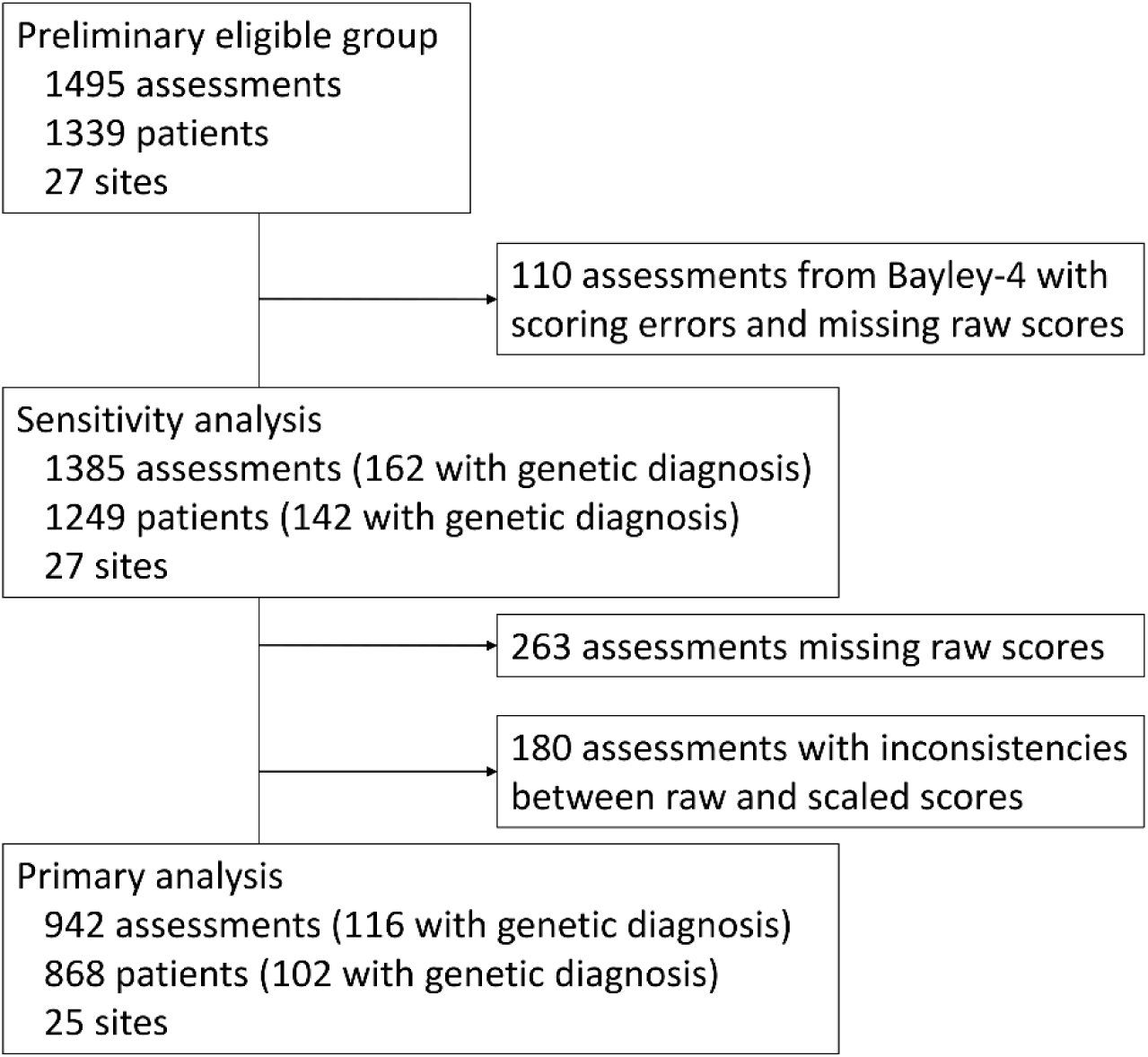
Flow diagram of patients from the Cardiac Neurodevelopmental Outcome Collaborative (CNOC) registry included in analyses.

As outlined in Table 1, 56.1% of patients were male, 55.5% were non-Hispanic White, preterm birth was present in 14.8%, and mean birthweight was 3.1 kg. The most common diagnoses were SVP (25.7%) and D-TGA (17.5%). Median age at surgery across the whole cohort was 20 days but varied significantly by diagnosis: 6 days for D-TGA, 9 for CoA, 125 for VSD, 107 for TOF, 8 for SVP, and 144 for AVSD. The median number and range of cardiac operations was 1 (1-5) and cardiac catheterizations was 0 (0-9). A history of cardiac arrest, stroke or seizures, and ECMO were all relatively uncommon (3.8-4.7%). The median post-operative length of stay at the first operation was 17 days for D-TGA, 24 for CoA, 8 for VSD, 14 for TOF, 35 for SVP, and 9.5 for AVSD. Among the 102 with a genetic diagnosis, 47 had trisomy 21, 28 had a 22Q11.2 deletion syndrome, and 27 had other genetic diagnoses.

**Table 1.** Birth, Sociodemographic, Medical, and Neurodevelopmental Characteristics of Patients by Genetic Diagnosis

|  | **Total**<br>N=868 | **No Genetic**<br>**Diagnosis**<br>N=766 | **Genetic Diagnosis**<br>N=102 |
| --- | --- | --- | --- |
| Total number of patients |  |  |  |
| *Birth characteristics* |  |  |  |
| Gestational age (wks) | 37.9 ± 2.0, n=836 | 38.0±1.9, n=735 | 37.2±2.3, n=101 |
| Preterm birth (<37 wks gestation) | 126/854 (14.8) | 99/752 (13.2) | 27/102 (26.5) |
| Birth weight (kg) | 3.1±0.6, n=753 | 3.1±0.6, n=673 | 2.8±0.7, n=80 |
| Head circumference (cm) | 33.2±2.1, n=412 | 33.3±2.1, n=368 | 32.2±2.1, n=44 |
| Sex: Female | 376/857 (43.9) | 330/757 (43.6) | 54/100 (54.0) |
| Male | 481 (56.1) | 427 (56.4) | 46 (46.0) |
| *Sociodemographic characteristics* |  |  |  |
| Race/ethnicity: Non-Hispanic Black | 85/695 (12.2) | 75/603 (12.4) | 10/92 (10.9) |
| Hispanic of any race | 142 (20.4) | 115 (19.1) | 27 (29.3) |
| Non-Hispanic White | 386 (55.5) | 337 (55.9) | 49 (53.3) |
| Other* | 82 (11.8) | 76 (12.6) | 6 (6.5) |
| Primary caregiver education: Graduate degree | 78/471 (16.6) | 70/407 (17.2) | 8/64 (12.5) |
| College degree | 163 (34.6) | 146 (35.9) | 17 (26.6) |
| Less than college | 230 (48.8) | 191 (46.9) | 39 (60.9) |
| Childhood Opportunity Index: Very High | 130/552 (23.6) | 120/476 (25.2) | 10/76 (13.2) |
| High | 121 (21.9) | 102 (21.4) | 19 (25.0) |
| Moderate | 117 (21.2) | 97 (20.4) | 20 (26.3) |
| Low | 97 (17.6) | 84 (17.6) | 13 (17.1) |
| Very Low | 87 (15.8) | 73 (15.3) | 14 (18.4) |
| *Medical characteristics* |  |  |  |
| Cardiac diagnosis: D-TGA | 152/868 (17.5) | 151/766 (19.7) | 1/102 (1.0) |
| CoA | 81 (9.3) | 69 (9.0) | 12 (11.8) |
| VSD | 87 (10.0) | 70 (9.1) | 17 (16.7) |
| TOF | 139 (16.0) | 118 (15.4) | 21 (20.6) |
| SVP | 223 (25.7) | 215 (28.1) | 8 (7.8) |
| AVSD | 40 (4.6) | 16 (2.1) | 24 (23.5) |
| Other | 146 (16.8) | 127 (16.6) | 19 (18.6) |
| Age at index operation (days) | 20 (6–127) | 13 (6–118) | 105 (34–151) |
| STAT category: 1 | 111/866 (12.8) | 90/764 (11.8) | 21/102 (20.6) |
| 2 | 135 (15.6) | 118 (15.4) | 17 (16.7) |
| 3 | 141 (16.3) | 113 (14.8) | 28 (27.5) |
| 4 | 319 (36.8) | 291 (38.1) | 28 (27.5) |
| 5 | 160 (18.5) | 152 (19.9) | 8 (7.8) |
| Events during index hospitalization |  |  |  |
| Cardiac arrest | 35/794 (4.4) | 31/698 (4.4) | 4/96 (4.2) |
| Stroke or seizure | 41/867 (4.7) | 38/765 (5.0) | 3/102 (2.9) |
| Extracorporeal membrane oxygenation | 33/867 (3.8) | 31/765 (4.1) | 2/102 (2.0) |
| Index operation hospital length of stay (days): ≤15 | 337/868 (38.8) | 296/766 (38.6) | 41/102 (40.2) |
| 16-30 | 220 (25.3) | 202 (26.4) | 18 (17.6) |
| 31-45 | 112 (12.9) | 99 (12.9) | 13 (12.7) |
| >45 | 199 (22.9) | 169 (22.1) | 30 (29.4) |
| Number of infant cardiac operations: 1 | 618/868 (71.2) | 535/766 (69.8) | 83/102 (81.4) |
| 2 | 215 (24.8) | 201 (26.2) | 14 (13.7) |
| 3 or more | 35 (4.0) | 30 (3.9) | 5 (4.9) |
| Number of infant catheterizations: 0 | 498/867 (57.4) | 425/765 (55.6) | 73/102 (71.6) |
| 1 | 246 (28.4) | 227 (29.7) | 19 (18.6) |
| 2 | 75 (8.7) | 67 (8.8) | 8 (7.8) |
| 3 or more | 48 (5.5) | 46 (6.0) | 2 (2.0) |
| *Neurodevelopmental characteristics* |  |  |  |
| Number of Bayley evaluations: 1 | 807/868 (93.0) | 717/766 (93.6) | 90/102 (88.2) |
| 2 | 50 (5.8) | 40 (5.2) | 10 (9.8) |
| 3 or more | 11 (1.3) | 9 (1.2) | 2 (2.0) |
| Total number of Bayley evaluations | N=942 | N=826 | N=116 |
| Age at testing (months) | 18.9 (15.3–22.9) | 18.9 (15.4–22.8) | 19.2 (14.9–24.6) |
| Bayley Version-III | 409/942 (43.4) | 363/826 (43.9) | 46/116 (39.7) |

| Version-4 | 533 (56.6) | 463 (56.1) | 70 (60.3) |
| --- | --- | --- | --- |
| Cognitive score | 88.8±18.1, n=929 | 91.1±17.2, n=817 | 72.3±15.2, n=115 |
| Language score | 86.2±19.1, n=874 | 88.2±18.7, n=780 | 72.1±16.1, n=113 |
| Motor score | 86.2±18.2, n=902 | 88.8±17.0, n=794 | 67.9±15.3, n=111 |

Values are mean±standard deviation, n/total (%), or median (interquartile range). AVSD indicates atrioventricular septal defect; Bayley, Bayley Scales of Infant Development; CoA, coarctation; D-TGA, dextro-transposition of the great arteries; STAT, Society of Thoracic Surgeons-European Association for Cardio-Thoracic Surgery; SVP, single ventricle physiology; TOF, tetralogy of Fallot; VSD, ventricular septal defect.
\* Other race/ethnicity in the total group includes multiracial (n=34), Asian (n=23), and other (n=25).

### Neurodevelopmental Outcomes

Across cardiac diagnostic groups, those with genetic diagnoses scored at least 1 SD (>15 points) lower on all Bayley indices than those without (*P*<0.001 for each, Table 1). Even amongst children without a genetic diagnosis, scores were significantly below the normative mean (100) for all three Bayley indices (*P*<0.001 for each). Figure 2 shows the consistent impact of genetics by type of genetic diagnosis. The lowest scores were observed in patients with trisomy 21, followed by 22Q11.2 deletion syndrome, and other genetic diagnoses.

**Figure 2.**
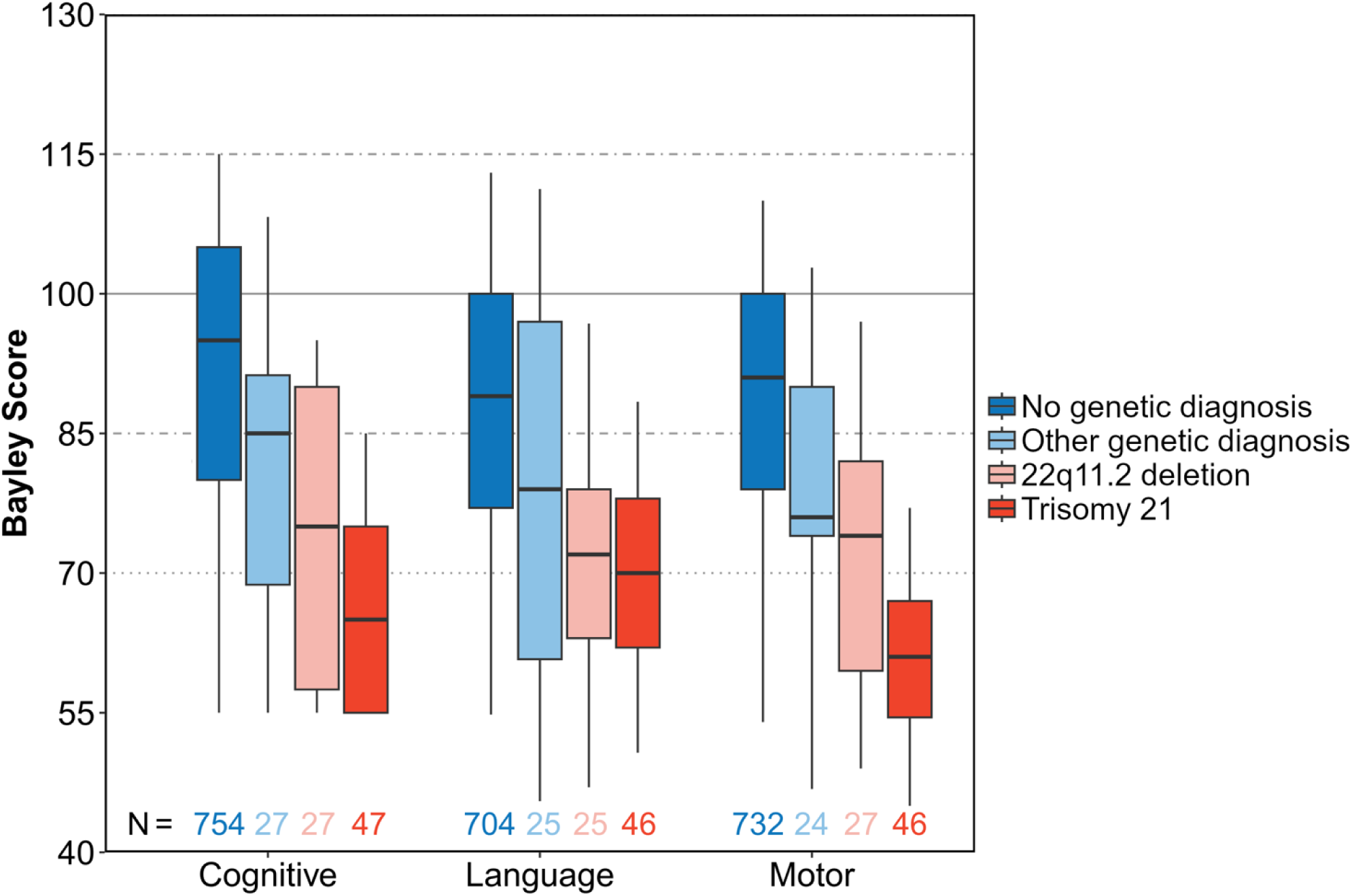
Boxplots of Bayley Cognitive, Language, and Motor scores by genetic diagnosis category. Patients with any genetic diagnosis scored lower in all indices than patients with no genetic diagnosis (*P*<0.001 for each). Lower and upper ends of the boxes represent the 25th and 75th percentiles, respectively; horizontal lines inside boxes represent the medians; whiskers extend to the 5th and 95th percentiles.

Bayley scores also varied by cardiac diagnosis and STAT category (Figure 3), separated by presence/absence of a genetic diagnosis. Patients with D-TGA without a genetic diagnosis exhibited the highest scores, with scores of 98.2±12.8, 94.4±16.3, and 96.8±12.1 (mean±SD) for Cognitive, Language, and Motor indices, respectively. Bayley scores were lower for STAT Category 5. Differences between children with or without a genetic diagnosis looked similar across cardiac diagnoses and STAT categories. Motor scores also varied by gestational age, birth weight, and head circumference (Figure 4); again, those with a genetic diagnosis had lower Bayley scores. In univariable analyses, Bayley scores also varied by primary caregiver education, hospital length of stay, and number of catheterizations (Figure 5).

**Figure 3.**
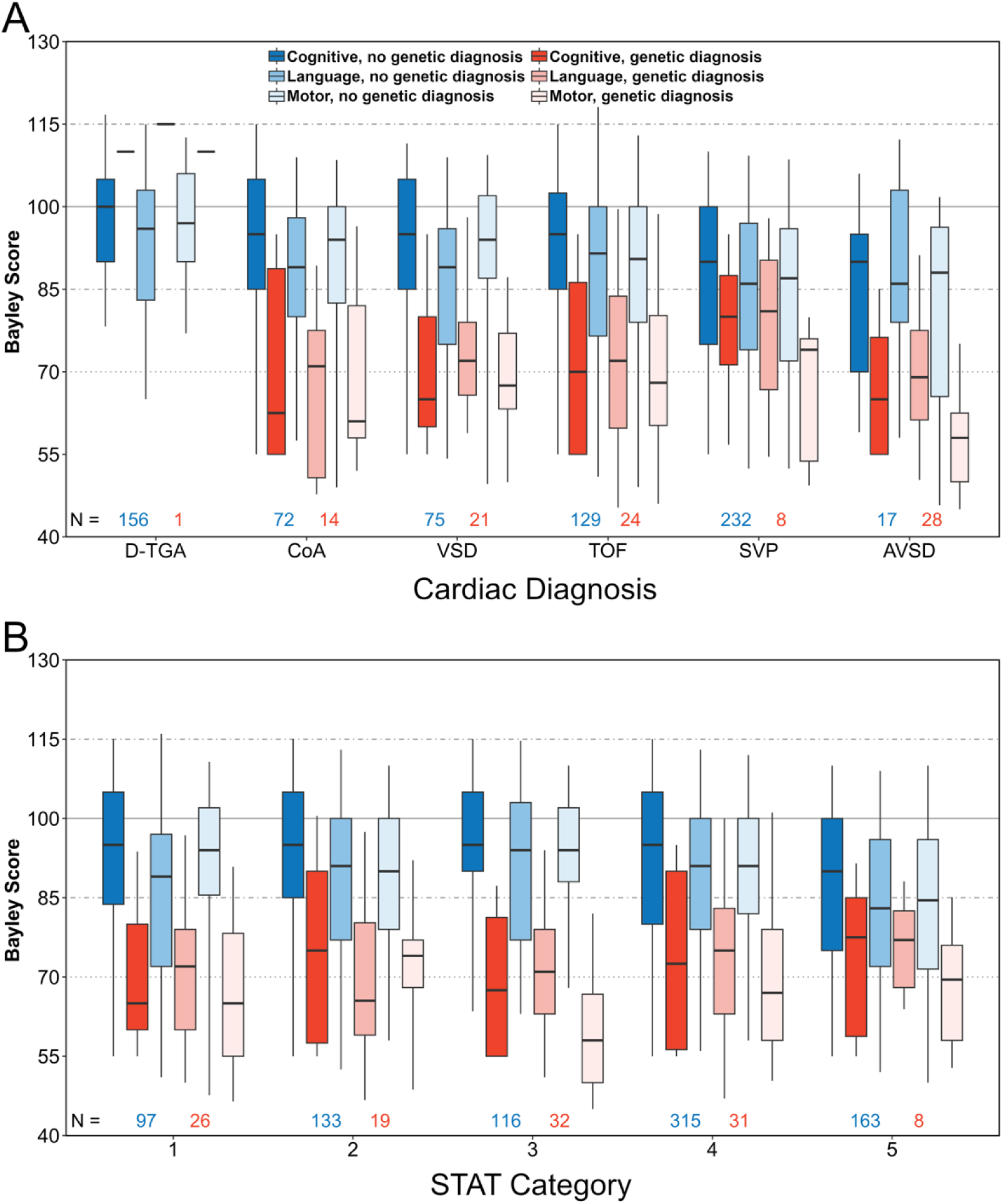
Boxplots of Bayley Cognitive, Language, and Motor scores by cardiac diagnosis (A) and STAT category (B) for patients with and without any genetic diagnosis. AVSD, atrioventricular septal defect; CoA, coarctation; D-TGA, transposition of the great arteries; STAT, The Society of Thoracic Surgeons-European Association for Cardio-Thoracic Surgery; SVP, single ventricle physiology; TOF, tetralogy of Fallot; VSD, ventricular septal defect. Lower and upper ends of the boxes represent the 25th and 75th percentiles, respectively; horizontal lines inside boxes represent the medians; whiskers extend to the 5th and 95th percentiles.

**Figure 4.**
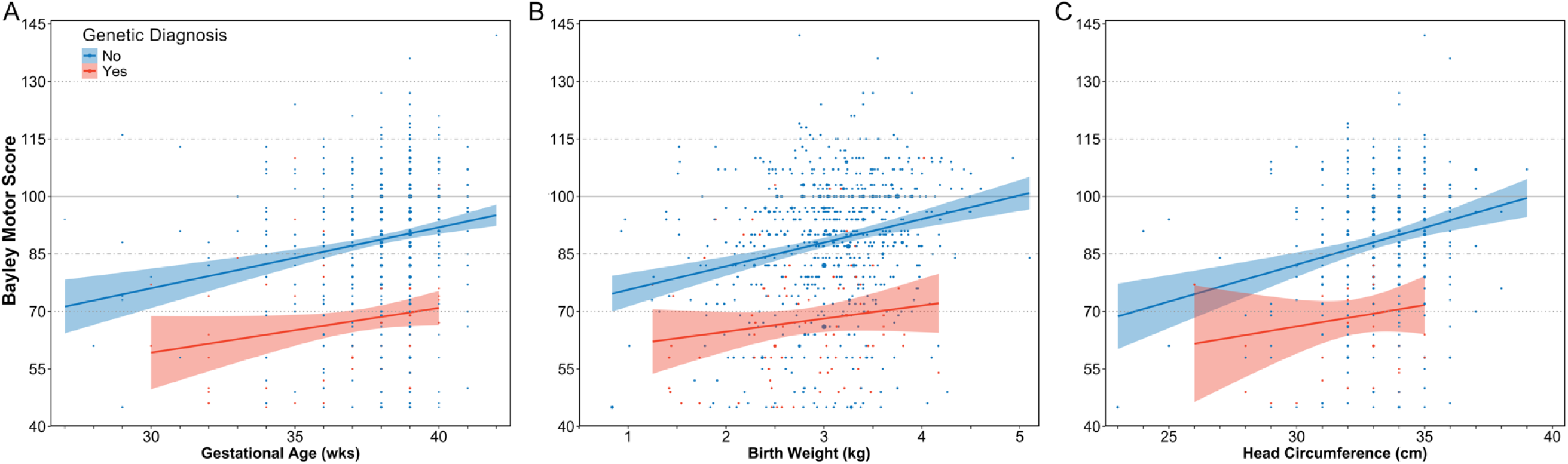
Scatterplots showing association of Bayley Motor scores with gestational age (A), birth weight (B), and head circumference (C) for patients with and without any genetic diagnosis. The size of the circles are proportional to the number of assessments at each point. Shaded areas represent pointwise 95% confidence limits around the fitted lines.

**Figure 5.**
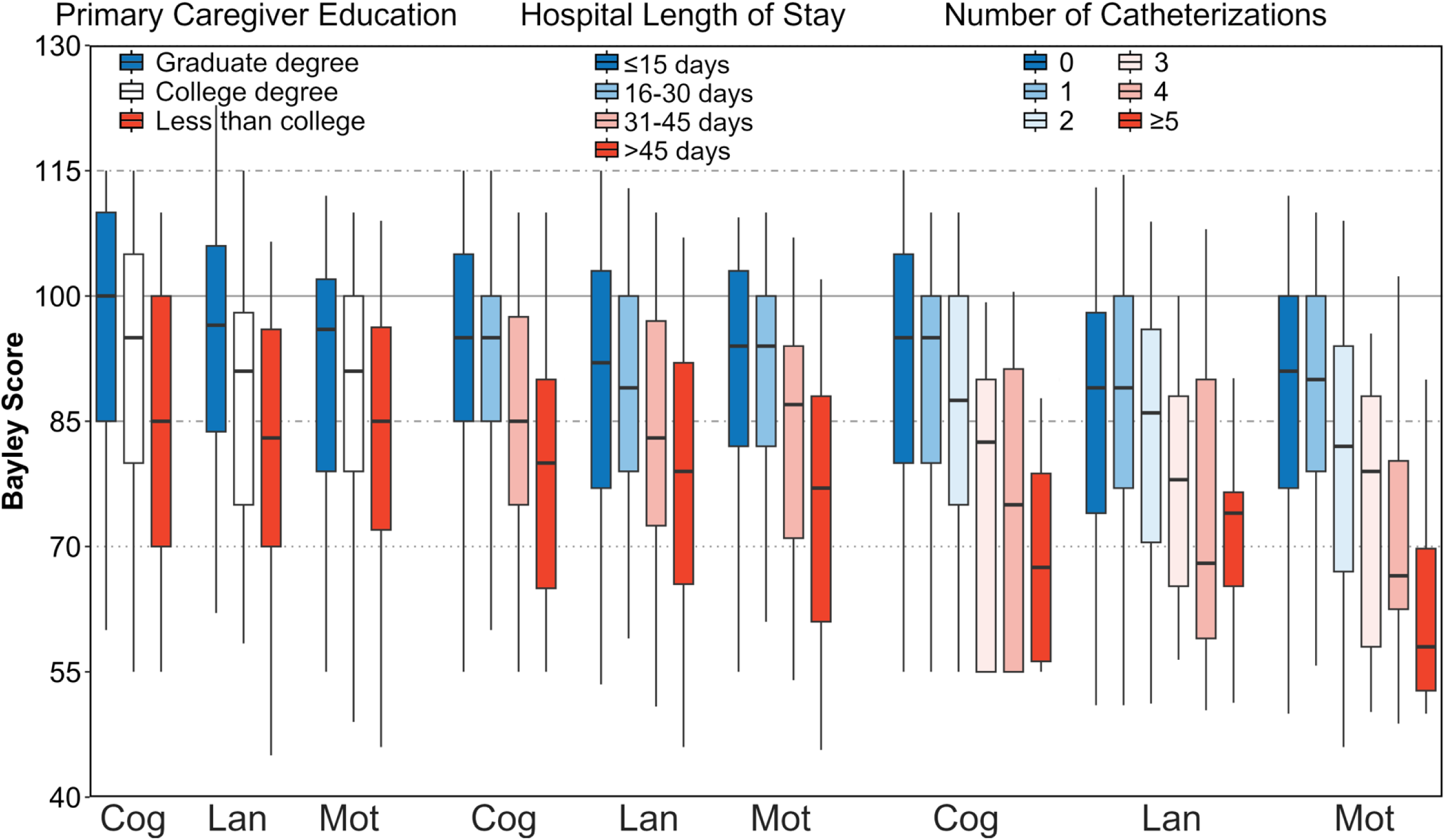
Boxplots of Bayley Cognitive, Language, and Motor scores by primary caregiver education, hospital length of stay, and number of catheterizations for patients without any genetic diagnosis. Cog, Cognitive; Lan, Language; Mot, Motor. Lower and upper ends of the boxes represent the 25th and 75th percentiles, respectively; horizontal lines inside boxes represent the medians; whiskers extend to the 5th and 95th percentiles.

### Multivariable Predictors of Neurodevelopmental Outcomes for Patients without a Genetic Diagnosis

The results of the multivariable model for patients without a genetic diagnosis are shown in Table 2. After adjusting for other variables, cardiac diagnosis was a significant predictor for Cognitive and Motor scores, with patients with D-TGA exhibiting the highest scores (Model 1). Lower birth weight, male sex, older age at first open-heart surgery, longer hospital length of stay, and number of infant catheterizations were independently associated with lower scores across all indices. A history of cardiac arrest, stroke or seizure, or ECMO were not predictive of ND outcomes for any models.

**Table 2.**
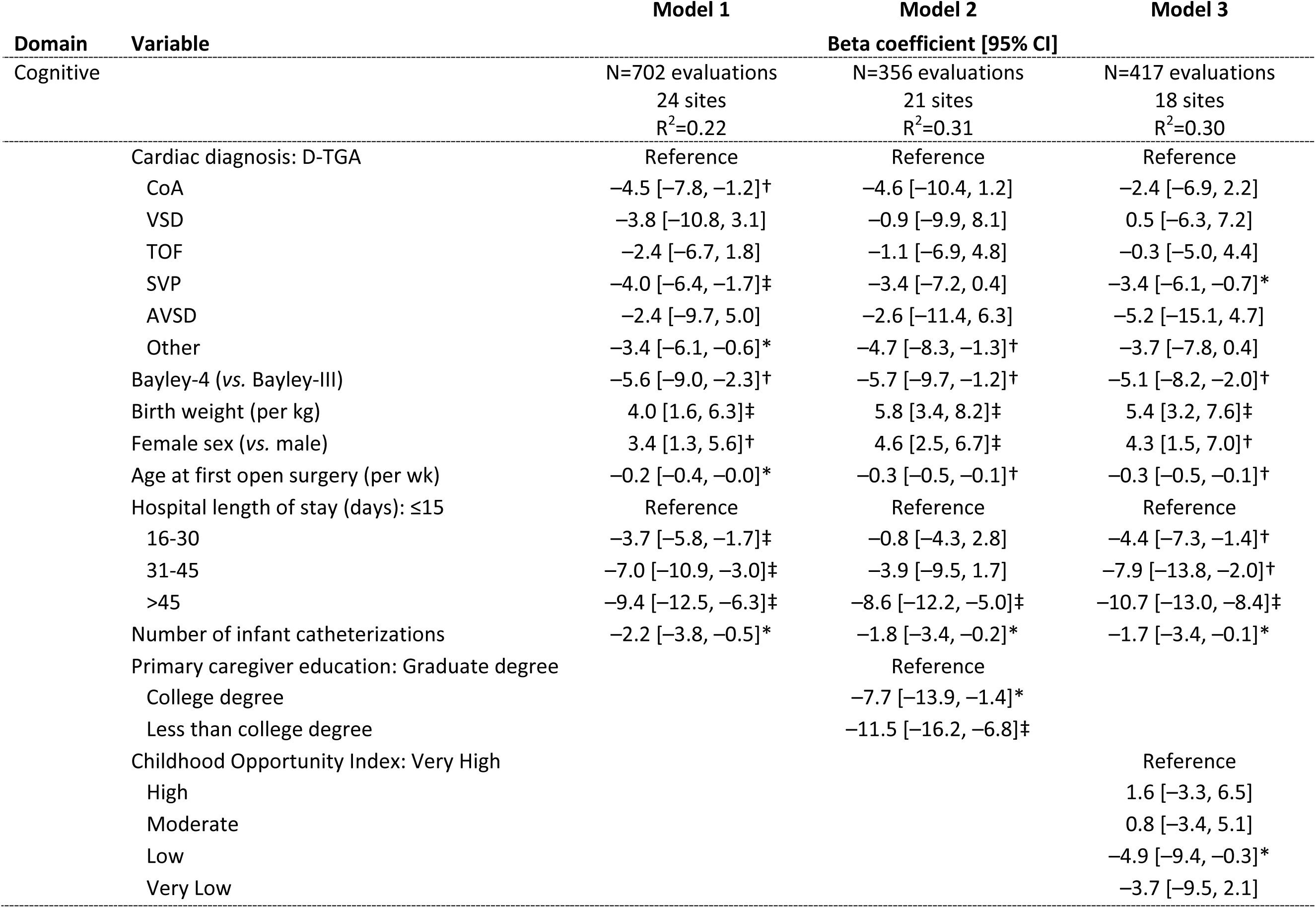
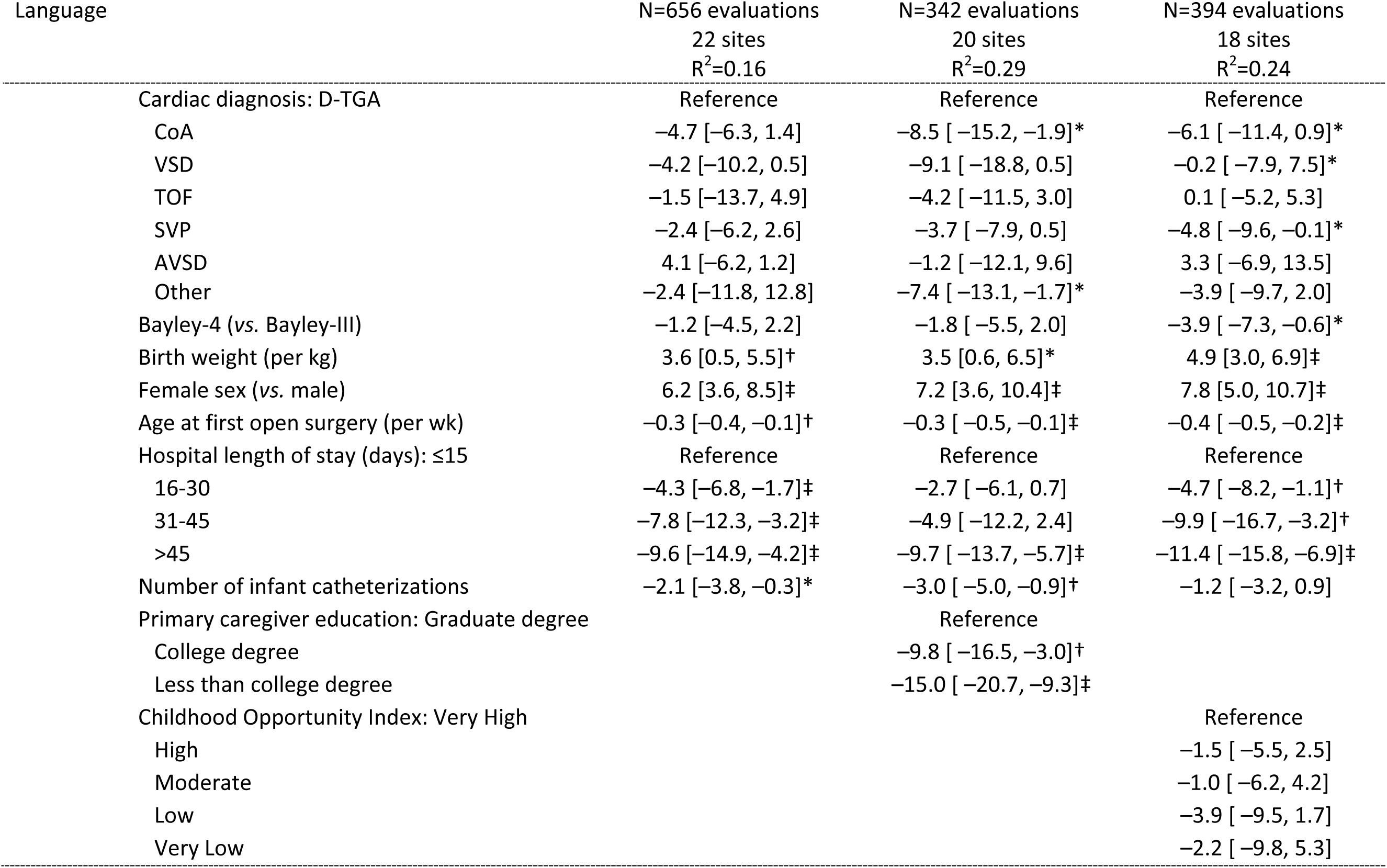
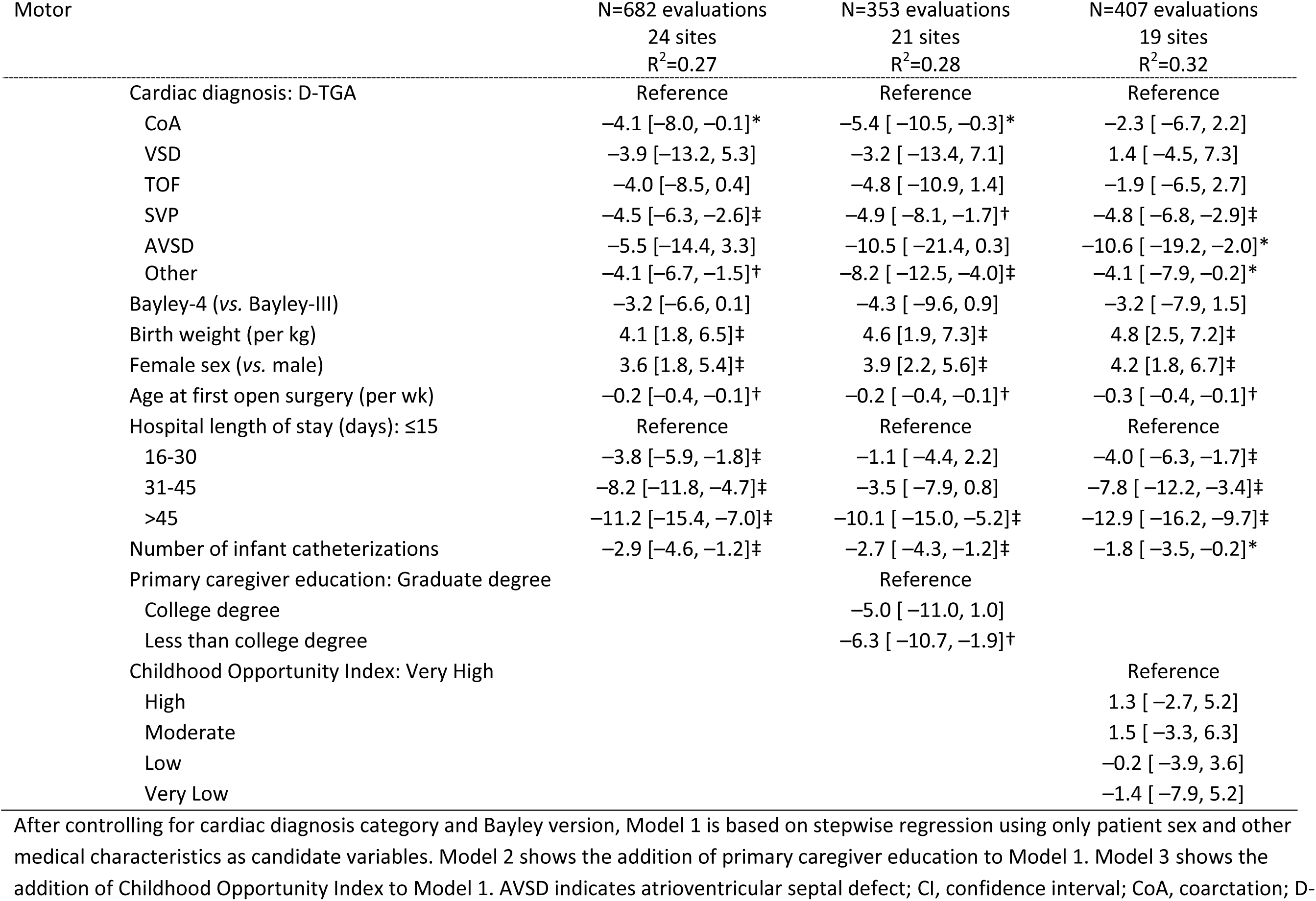
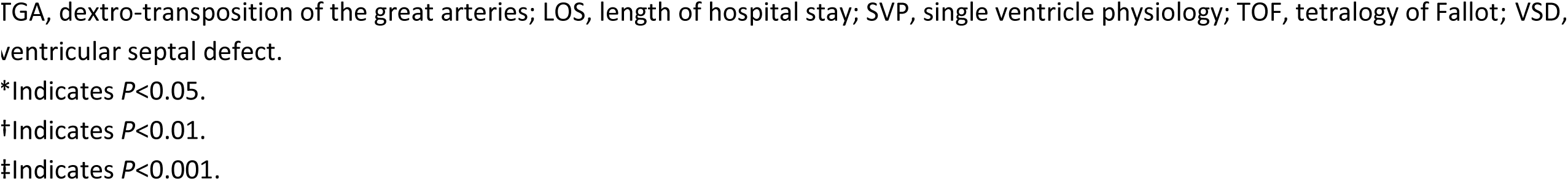
Multivariable Predictors of Neurodevelopmental Outcomes on the Bayley Scales of Infant Development

When the models included primary caregiver education (Model 2), the same medical factors remained statistically significant, while primary caregiver education was also a strong predictor of outcome. The largest differences were found for the Language index, where children of primary caregivers with higher education scored higher than those without a college degree (15.0 points higher for those with a graduate degree and 9.8 points higher for college education). COI was predictive of the Cognitive index, with the low COI group having the lowest scores. COI was not predictive of Language or Motor indices (Model 3). Regression analyses using the sensitivity analysis dataset (Figure 1) of patients without a genetic diagnosis included 1223 assessments representing 1107 unique patients from 27 sites, comprising 1050 assessments for the Cognitive index, 995 for Language, and 986 for Motor. These analyses yielded similar inferences for Models 1, 2, and 3, with D-TGA patients exhibiting significantly higher Language, Cognitive, and Motor scores, and COI also significantly predictive of Language and Motor indices.

## Discussion

This initial analysis of infant and toddler ND data from the CNOC registry supports the hypothesis that cardiac diagnosis is predictive of early ND outcomes. Those with D-TGA exhibited significantly higher scores on the Bayley than those with other forms of CHD, while those with AVSD and SVP exhibited the lowest scores amongst the groups. Our findings are consistent with prior studies demonstrating that the presence of a genetic diagnosis is a strong predictor of ND outcomes across all cardiac diagnoses.^5^ Beyond genetic diagnosis, other significant medical and social risk factors for adverse ND outcomes in patients with CHD without a genetic diagnosis included lower birthweight, male sex, older age at initial surgery, longer hospital length of stay, more cardiac catheterizations, and lower primary caregiver education. Lower COI scores were also a predictor of lower Cognitive scores. Previously identified medical risk factors that were not significantly associated with ND outcomes in this young cohort included cardiac arrest, a history of stroke or seizures, and ECMO.

Findings from this CNOC registry cohort need to be considered in the context of prior studies, including differences in the approach to classifying cardiac diagnoses, variation in outcomes associated with Bayley edition, age at ND assessment, and effects of surgical era. Broadly speaking, prior studies have reported more favorable early ND outcomes in patients undergoing biventricular repairs than those requiring single ventricle palliation. Reductions of half a standard deviation or more on the Bayley are consistently reported for children with SVP. The International Cardiac Collaborative on Neurodevelopment Investigators reported Bayley-II scores for a large heterogeneous group of patients with CHD who were tested at 14.5±3.7 months of age (n=1770 from 22 institutions over a 13-year period).^13^ Patients were divided into four groups of cardiac diagnoses based whether they had undergone a biventricular repair versus single ventricle palliation, with or without aortic arch obstruction. Those with univentricular hearts with arch obstruction scored 6.3-13.6 points lower than those with biventricular repairs without arch obstruction, while those with univentricular hearts without arch obstruction scored 3.1-9.1 points lower. Bayley-III assessment at 2 years of age in a cohort enrolled at The Royal Children’s Hospital, Melbourne and Starship Children’s Hospital, Auckland, revealed that children with SVP without arch obstruction exhibited the lowest scores, while the group with two ventricles with arch obstruction composed primarily of patients with CoA performed better than those with D-TGA.^26^ In a cohort of infants with either D-TGA or SVP enrolled at the University of California San Francisco and the University of British Columbia, those with SVP scored an average 12 points lower than those with D-TGA on the Bayley-II.^27^ This same group has also found that children with other forms of CHD, including left- or right-sided obstructive lesions, anomalous pulmonary venous return, and truncus arteriosus had lower Bayley-II Cognitive scores compared to those with HLHS and D-TGA, though worse ND outcomes in this group were driven by those with genetic disorders.^28^

Compared with the findings of the Boston Circulatory Arrest Trial cohort, the motor scores we observed in infants with D-TGA suggest there may have been an improvement in outcomes for children with D-TGA over the past two decades. The Boston Circulatory Arrest Trial enrolled newborns from 1988-1992 and found a mean score for psychomotor development of 95.1±15.5 at one year using the original Bayley scale. This apparent improvement in motor outcomes could reflect advances in surgical techniques and perioperative management occurring during the three decades since this seminal study, while some variation in Bayley scores may reflect version-specific bias. It is important to note that despite the average abilities for early cognitive development reported by the Boston Circulatory Arrest Trial, subsequent follow-up at school age and later revealed a high prevalence of neurocognitive and psychosocial concerns in this cohort, emphasizing that average scores during infancy and toddler years should not be regarded as entirely reassuring with respect to subsequent development.^16,17^ However, it is interesting to consider possible improvements in ND outcomes in D-TGA in relation to our finding that older age at surgery was associated with lower scores on the Bayley. One prior study demonstrated that brain growth and language development were adversely impacted by delayed surgery for D-TGA, which was more common in previous eras.^29^ The adverse impact of unrepaired CHD physiology on infant growth and nutrition as a potential risk factor for impaired brain development has been emphasized by the updated American Heart Association ND guideline.^2^ With respect to prenatal brain development, it is also interesting to consider that patients with D-TGA are spared some of the more profound fetal circulatory disturbances and impairment of fetal brain growth seen in more severe forms of CHD such as SVP.^30^

While this analysis did not find an association between stroke or seizures and Bayley scores, many centers do not routinely survey infants with post-operative electroencephalography or perform routine brain magnetic resonance imaging (MRI) in the perioperative period. Moderate-to-severe white matter injury on brain MRI has previously been shown to be a significant predictor of Bayley-II Psychomotor and Mental Development Indices, while stroke was unrelated.^27^ Other studies with mixed samples of CHD found that abnormal neuroimaging was associated with a clinical neurological exam whereas abnormal neuroimaging was not related to findings on standardized neurological outcome measures.^31,32^ The initial findings from the CNOC registry prompt us to reconsider how to conceptualize risk specific to CHD subtype. Attention has so far focused on the severity of the heart disease and complexity of the index operation. However, while patients with D-TGA are considered medically higher risk than those with less profound cardiac malformations such as CoA or VSD, ND outcomes for D-TGA may be better, at least at early follow-up. Similarly, the current analysis of the CNOC data suggest that the use of a CHD classification system based on the presence of biventricular versus univentricular physiology with or without aortic arch obstruction results in a wide variety of cardiovascular physiologies and timing of surgery being grouped together that may reduce the prognostication possible with more specific diagnoses. One example is the somewhat unexpected finding that among patients without a genetic diagnosis, those with AVSD exhibited lower ND outcomes than patients with other cardiac diagnoses, including SVP.

Prolonged hospital length of stay was an important predictor of outcome in our analysis of the CNOC registry data. Longer hospitalizations have been associated with adverse ND outcomes in the setting of CHD in multiple studies.^33,34^ The basis of this association is likely related to a range of factors including innate patient factors and treatment effects that reflect medical complexity, also indicated by the association we observed between ND outcomes and number of cardiac catheterizations. Similarly, in our univariable analysis, patients in STAT category 5 from the CNOC registry exhibited lower ND scores. A complex course in hospital may interfere with brain development through a number of mechanisms including abnormal cerebrovascular physiology, brain injury, decreased developmental interactions due to medical fragility, and poor nutrition. Our data therefore reinforce the importance of an approach to inpatient care that promotes optimal neurodevelopment and minimizes the impact of intensive care on developmental trajectory.^35,36^

The impact of genetic variation on brain development that has previously been identified in the setting of CHD is reinforced by the findings from the CNOC registry. Of note, in CHD patients with a genetic diagnosis, there were fewer differences in ND outcomes between different cardiac diagnoses than were present in patients without a genetic diagnosis. However, we did observe differences in ND outcomes between different genetic diagnoses, whereby patients with trisomy 21 scored lower than those with 22Q11.2 deletion syndrome while those with other diagnoses scored higher than those with trisomy 21 and 22Q11.2 deletion syndrome. In the future, the application of deeper genomic analysis techniques in CNOC registry patients may also reveal associations between genetic variation and ND outcomes in patients without a genetic diagnosis.

This initial analysis of the registry also supports the findings of other studies that have identified the importance of social drivers of health on ND outcomes.^8,9^ In keeping with our findings, prior studies have shown that higher parent education is associated with improved early developmental skills, as well as fewer behavioral and emotional concerns.^13,18,37^ Other studies in children with CHD have explored neighborhood level factors and shown that higher median household income, lower percentage below poverty, and lower percentage unemployment rate are associated with higher infant and toddler abilities.^19,38^ While our current findings showed only a modest association for cognitive outcomes with COI, this may be attributable to limited zip code data currently available through the CNOC registry.

There are several limitations of this study. The data are contributed through the clinical CNOC registry, whereby some variables cannot be entered by some sites, including zip code. The size of our dataset was further limited by our conservative approach to excluding patients for whom the Bayley raw scores were not available. It is recognized that clinical registry data may be subject to patient heterogeneity and practice variation by site. Selection bias is also an important limitation of this study as attendance in cardiac ND programs is associated with risk factors that may impact development, including some social drivers of health, as we previously identified from CNOC registry data.^22^ For example, in this prior study only 29% of eligible CHD patients returned for follow-up, while longer distance from hospital was associated with lower follow-up rates, emphasizing the importance of addressing social drivers of health to optimize ND outcomes. Despite these potential shortcomings of registry research, our findings align with the 2024 Scientific Statement of the American Heart Association, which recommends serial developmental evaluation for all children undergoing cardiac surgery with cardiopulmonary bypass during infancy to detect ND issues early thus facilitating timely intervention.^2^ Future directions for this research include longitudinal ND follow-up of this cohort. With the anticipated increase in sample size, analysis of medical risk factors and social drivers of health within individual CHD lesions will be possible. The registry data may also provide a more data-driven and therefore equitable approach to risk stratification in the setting of resource limited health care settings. Although we recognize the current limitations of our ability to accurately predict ND outcomes in this population, we have demonstrated that conventional approaches to stratifying participants by cardiac diagnosis alone is inappropriate.

In conclusion, analysis of this initial but sizeable sample collected by the CNOC registry confirms that cardiac diagnosis is associated with early ND outcomes, with improved outcomes now being observed in participants with D-TGA in toddlerhood. Previously identified medical risk factors including preterm birth, low birthweight, and prolonged hospitalization were found to confer an increased risk of adverse ND outcomes, while older age at surgery and number of cardiac catheterizations may also pose a risk to neurodevelopment. The presence of trisomy 21 and 22Q11.2 deletion syndrome remain the most important risk factors for ND delay, while previously identified social drivers of health such as parental education and COI were also significant. The finding of lower scores on ND testing in children with somewhat less severe forms of CHD such as CoA, VSD, TOF, and AVSD compared with D-TGA emphasizes the importance of follow-up for all infants undergoing infant heart surgery.

## Data Availability

Data available upon request

## Acknowledgements

None

## Sources of funding

Cardiac Neurodevelopment Outcome Collaborative

## Disclosures

None

## Non-standard Abbreviations and Acronyms

AVSD: atrioventricular septal defect
CHD: congenital heart disease
CNOC: Cardiac Neurodevelopmental Outcome Collaborative
COA: coarctation of the aorta
COI: Child Opportunity Index
D-TGA: dextro-transposition of the great arteries
ECMO: extracorporeal membrane oxygenation
HLHS: hypoplastic left heart syndrome
MRI: magnetic resonance imaging
ND: neurodevelopmental
PC^4^: Pediatric Cardiac Critical Care Consortium
STAT: Society of Thoracic Surgeons-European Association for Cardio-Thoracic Surgery
SVP: single ventricle physiology
TOF: tetralogy of Fallot
VSD: ventricular septal defect

## Notes

### Competing Interest Statement

The authors have declared no competing interest.

### Funding Statement

No external funding recieved

### Author Declarations

University of Michigan Institutional Review Board

